# Impact of COVID-19 on deaths from respiratory diseases: Panel Data evidence from Chile

**DOI:** 10.1101/2021.06.20.21259216

**Authors:** Claudia Barría-Sandoval, Angie Mendez, Guillermo Ferreira, María Cecilia Toffoletto

## Abstract

The COVID-19 pandemic has left other pathologies commonly present in the population in a secondary context. Therefore, it is necessary to study the evolution of these diseases in the presence or absence of COVID-19.

**Objectives:** The present study had the following objectives: 1. To evaluate the relationship between the COVID-19 epidemic and the possible decrease in deaths from respiratory diseases in Chile. 2 Study the relationship between meteorological variables and severity of COVID-19 with respect to deaths from respiratory diseases from January 2018 to February 2021.

**Methods:** The variable number of deaths due to respiratory diseases in Chile was analyzed considering the monthly records of meteorological variables (temperature, precipitation and humidity) in each Region of Chile and severity of COVID-19, to evaluate the mortality trend before and after the pandemic. For this, different Non-Observable Heterogeneity Models for Panel Data were used

**Results:** Our findings show that among the variables that affect the mortality rate from respiratory diseases, there are the number of deaths from COVID-19 that has a negative effect, the number of patients with COVID-19 in intensive care unit (ICU) that has a positive effect and the minimum temperature with a negative effect. These results are supported by the application of the panel regression with one-way random-effects models.

**Conclusion:** This study revealed that there is an unexpected reduction in deaths from respiratory diseases in Chile in the post-pandemic period. Therefore, it can be concluded that this variable decreased with the appearance of the COVID-19 pandemic.

## Introduction

In December 2019, the People’s Republic of China reported a cluster of pneumonia cases of unknown etiology, subsequently identified as a new coronavirus by the Chinese Center for Disease Control and Prevention. Later time, in February 2020, the World Health Organization (WHO) named the disease COVID-19, short for “coronavirus disease 2019” and, in March of the same year, declared an international public health emergency for its rapid community, regional and global spread. From the confirmation of the first COVID-19 cases to March 2021, 116.736.437 confirmed cases were reported, including 2.593.285 deaths worldwide, of which 45 % of confirmed cases and 48 % of deaths were contributed by the Region of the Americas. Specifically, South America contributed with a higher proportion of deaths (85 %) of the total, surpassing North America (14.5 %) [1].

In March 2021, the Ministry of Health of Chile (MINSAL for the Spanish acronym for Ministry of Health) published in its epidemiological report [2], that up to that date a total of 1.018.677 cases (confirmed and probable) and a total of 28.756 deaths from COVID-19 had been registered. Another report [3] also prepared by the MINSAL, describes the analysis of the causes of deaths from COVID-19, where Chile applied rules for coding mortality and simulating the profile of deaths from COVID-19, not considering the existence of the virus as a cause death in medical certification. This evidenced that respiratory causes would have the first place in deaths in the country, observing that deaths from Influenza and Pneumonia represent 47 %, and deaths from Other Diseases of the Respiratory System represent 5 %, displacing even circulatory diseases and tumors, which in previous years occupied the first places in the ranking of deaths in the country.

In Chile, in 2018 (pre-pandemic) respiratory diseases ranked third in the national ranking of cause of death, only surpassed by diseases of the circulatory system and cancer; observing a total of 12.228 deaths from this cause [4]. However, as indicated in the previous paragraph, this situation changed with the arrival of COVID-19 in the country.

A recent study published by [5] reveals that 545 million people currently live with respiratory diseases (Rd hereafter), which represents 7.4 % of the world population and that 3.9 million people die each year from Rd, which constitutes a great health burden, due to its severity and mortality figures worldwide. The associated healthcare costs are an increasing burden on the economies of all countries, and if the loss of family or caregiver productivity of people with respiratory diseases is considered, the cost to society is much higher [6]. Likewise, at the national level in people over 65 years of age, Rd are the second cause of hospital discharge [7]. The previous discussion reflects the need to carry out a study on Rd in the pre and post pandemic period so that health entities are aware of a quantitative measure of how COVID-19 has affected the statistics on deaths from respiratory diseases, in such a way that this information can contribute to the effective distribution of health resources.

There are several studies that have addressed the health impact relating mortality from respiratory diseases and mortality from COVID-19. In this context, a Brazilian study made comparisons between deaths from respiratory diseases and deaths from COVID-19 with a time series statistical methodology for monthly registry data, in order to describe the peak of mortality during the summer seasons. This study revealed a latitudinal gradient in the country, with 27 peak deaths occurring in April in most of the northern and northeastern states, and gradually later in the southernmost states, with peak mortality in June and July (season of cold) in the Southeast and South regions, respectively [8].

On the other hand, a Spanish study used four regression models to explore the associations of the average daily temperature and air quality (PM 2.5) with the new daily cases of COVID-19 in the four main regions of Spain (Castilla y León, Castilla-La Mancha, Catalonia and Madrid) [9]. Among these models are, Panel Regression Models, Quantile Regression Model, Clustered OLS and Fixed Effects Regression Model known as Individual Heterogeneity Models. Recently, a study used Individual Heterogeneity Models to examine the effect between COVID-19 using the variables confirmed cases and deaths, which reflect the severity of the pandemic, and the meteorological factors and the air pollutant (PM 2.5), in six countries in South Asia. The authors have concluded that high temperature and humidity increase the transmission of COVID-19 infections and that it can also be applied to regions with higher transmission rates, where the minimum temperature is mostly above 21◦ C and the Humidity hovers around 80 % for months. Additionally, the air pollutant (PM 2.5) exhibits significant negative and positive effects on the number of confirmed COVID-19 cases [10]. Other investigations have also used Individual Heterogeneity Models as statistical methodology to examine the relationship between meteorological variables and environmental pollution with respect to confirmed cases of COVID-19 and mortality, [11],[12] and their references.

Another study evaluated the co-infection of COVID-19 in the presence of other respiratory viruses, analyzing the statistically significant correlations of the variables. In particular, [13] evaluated the presence of influenza A/B virus, human metapneumovirus, bocavirus, adenovirus, respiratory syncytial virus, and parainfluenza virus in 105 patients who died from COVID-19; finding co-infection with influenza virus in 22.3 %, respiratory syncytial virus and bocavirus in 9.7 %, parainfluenza virus in 3.9 %, human metapneumovirus in 2.9 % and finally adenovirus in 1.9 % of COVID-19 positive dead cases. In the same line [14] suggested higher rates of co-infection between SARS-CoV-2 and other respiratory pathogens. There are also studies that have explored the possibility that COVID-19 has affected the seasonal trend of other viruses, for example, the circulation of influenza has been decreased see [15]. However, it circulates together with COVID-19, being able to generate a greater impact on the evolution and management of respiratory diseases [16]. An important aspect that has aroused great interest among different scientific groups is the relationship between influenza and the COVID-19 outbreak. In this context, a study [17] analyzed whether influenza vaccination minimizes the spread of COVID-19 in the Italian population aged 65 to 48 years or older; concluding that the influenza vaccination coverage rate in people 65 years of age and older was associated with reduced spread and less severe clinical expression of COVID-19. The reader can deepen this topic by reviewing the studies carried out in Europe and the USA. [18],[19] and [20].

Among the various topics about the impact of COVID-19 and its relationship with respiratory diseases, there are studies that compared the rates of transmissibility, hospitalization and mortality of COVID-19 with respect to other pathogens that cause respiratory diseases [21]. The review of the research mentioned in the previous paragraphs highlights the effects that the COVID-19 pandemic has had on the health of the population. So much so, that other pathologies that cause great damage or deterioration of life and even that can cause death have been neglected. It is in this context, it seems necessary to study the effect of the COVID-19 pandemic outbreak that has had on deaths from respiratory diseases in Chile, considering the regional effect and climatic variables that predominate throughout the country.

In this study we focus on respiratory diseases, classified in J00-J99 according to the International Classification of Diseases (ICD 10); and that constitute those pathologies of the respiratory tract and other structures of the lung, associated with the main causes of morbidity and mortality worldwide [22]. In particular, this research proposes to quantify the relationship between deaths from respiratory diseases with the pandemic generated by COVID-19 and with meteorological variables in Chile. To do this, we generate a data panel with monthly records from different regions of Chile from January 2018 to February 2021 and Individual Heterogeneity Models are proposed, such as; Fixed and Random Effects Regression Models. To the best of our knowledge, this of the few studies to use panel data regression analysis to determine the effect of deaths from respiratory diseases and COVID-19 outbreak. The objective of this work is to provide an empirical study to determine the variables that have affected mortality from respiratory diseases in Chile in the period of the COVID-19 pandemic. In this way, it is intended to contribute useful statistical information for government authorities on the evolution of deaths from respiratory diseases in Chile with respect to the COVID-19 pandemic and meteorological variables. Another aspect that we must highlight from this study is that once COVID-19 is controlled in the population, chronic diseases that have been neglected in this period will reappear with force, and therefore, have information on the effect of COVID-19 on these chronic diseases is crucial to effectively allocate both available human and financial resources. In this context, this study has focused on deaths from respiratory diseases, without prejudice that it can be extended to other chronic diseases which must be continuously monitored by the Health authorities.

The rest of the article is organized as follows: Section 1 introduces the data, defines the variables, along with the empirical models; Section 2 reports the empirical results. Section 3 presents the strengths and weaknesses of this study through a brief discussion, and the last section concludes the study.

## 1. Materials and methods

### Data Sources and the Definition of Variables

To carry out this study, the monthly records of the variables (Table 1) were considered for 14 regions of Chile, from January 2018 to February 2021. These variables include the data on the number of deaths from respiratory diseases obtained from the Ministry of Health https://deis.minsal.cl/, whereas the data on the variables regarding the severity of COVId-19 were obtained from the Ministry of Science and Technology https://www.minciencia.gob.cl/covid19. Finally, the data of the temperature and precipitation variables were obtained from the updated report of the Chilean Meteorological Directorate DGAC, which are available on the website https://climatologia.meteochile.gob.cl. With this information, a balanced data panel was generated with a total of 14 regions and 38 temporal observations. Figure (1) shows a map of Chile with the regions where data recording was possible (gray color). In the regions highlighted in white, it was not possible to build the observation panel since there are no meteorological stations in these geographical areas that allow obtaining information on temperature and precipitation. Table (1) describes the variables used in this study. In particular, the dependent variable is the number of deaths from respiratory diseases (DRd). In addition, to investigate the impact of COVID-19 on the number of DRd, we consider three variables, namely: the monthly number of confirmed deaths from COVID-19 (CD), the incidence rate (IC), and the monthly number of patients in the intensive care unit diagnosed with COVID-19 (ICU).

**Figura 1.**
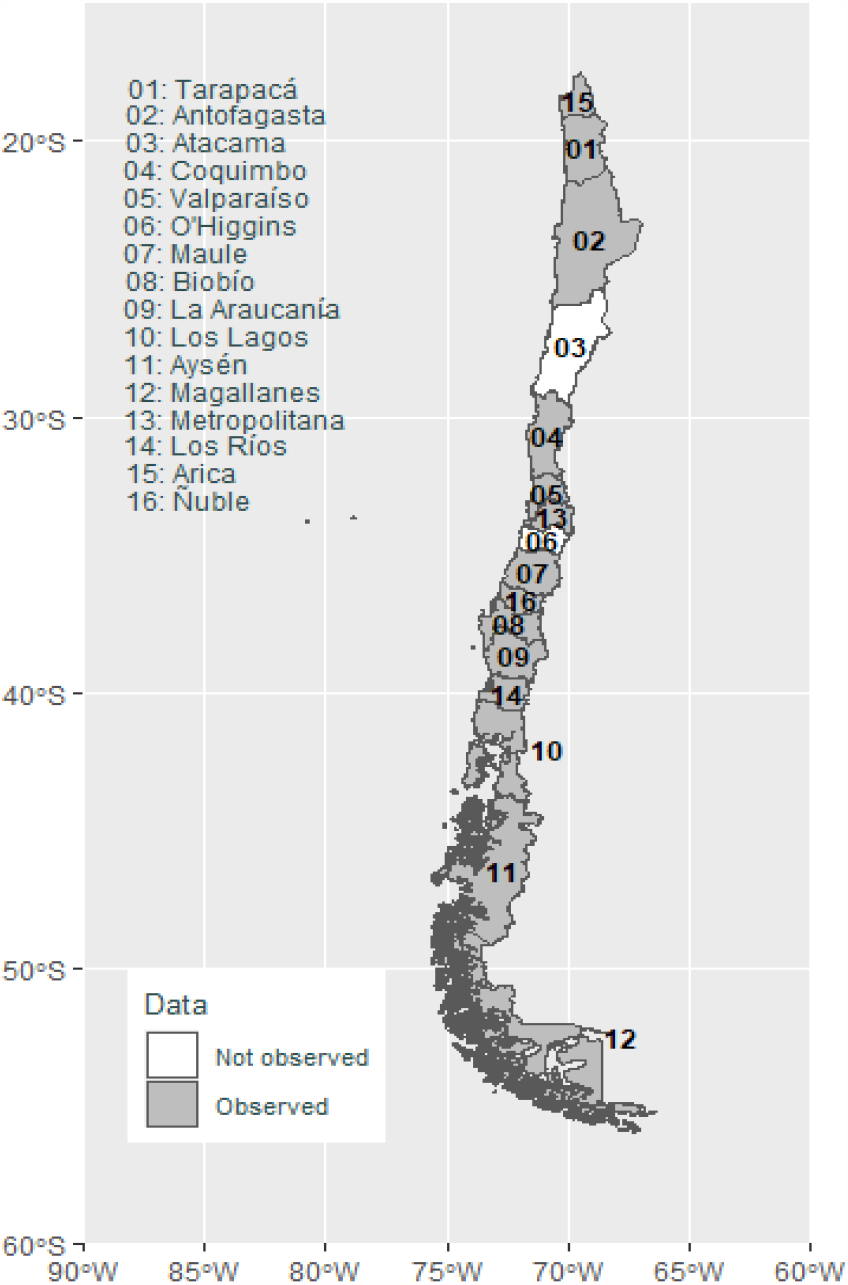
Chile’ regiones. Dataset

**Cuadro 1.**
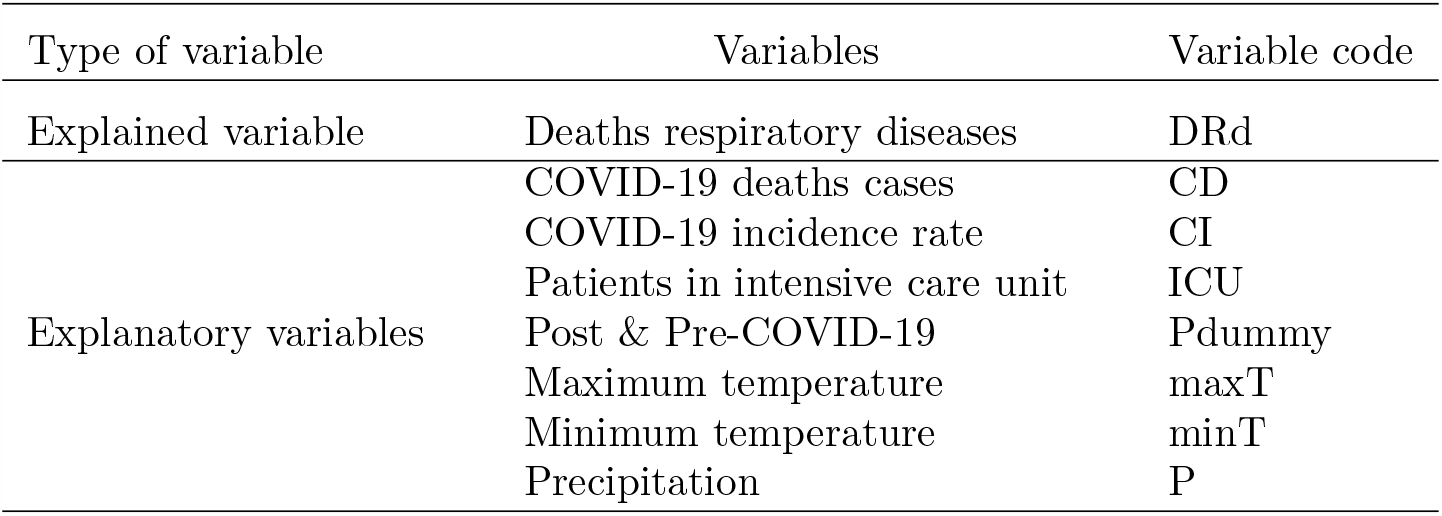
Varibales under study

### Empirical Models

To quantify the impact that the COVID-19 pandemic has had on the number of DRd in Chile, we used a panel data regression. In this regard, we can point out that the panel data models take into account time and regions simultaneously, while other models have the limitation of only expressing these heterogeneities across units or over time. Furthermore, panel data models are better at capturing the heterogeneity involved in both cross-sectional units and time dimensions, to reduce estimation bias and multicollinearity, and are better suited for studying the dynamics of change and complex behavioral models [23] and [24]. In this study, we consider two types of panel data models. The first, denoted by Model 1, is a two-way fixed or random effects model, that is, this model captures the individual and temporal effect of the data. While the second model, called Model 2, is a one-way random or fixed effects model, which only considers the temporal effect of the data.

▪ Two-way fixed-effects model First, we consider a two-way fixed-effects model with all the explanatory variables as follows: Model 1

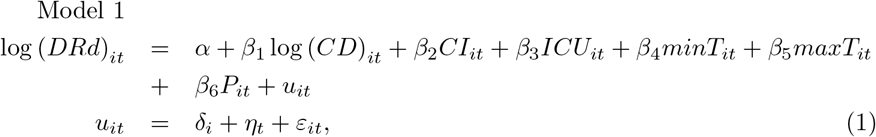

where *DRd* denotes the number of deaths from respiratory diseases, *CD* is the number of monthly deaths from COVID-19, the log (*CD*) is with respect to the observations obtained at the beginning of the pandemic, that is, observed from March 2020, *CI* is the incidence rate, which is the number of monthly confirmed cases divided by the gross permanent resident population (number confirmed/population (10.000 per unit)), *maxT minT* denotes temperature (measured in Celsius), *P* denotes precipitation (measured in hPa), *ICU* is the number of COVID-19 patients in intensive care, *i* denotes the region, *t* denotes the period under study, the *δ*_*i*_ and *η*_*t*_ are the region and time fixed effects, respectively, and *ε*_*it*_ denotes the error term. In the case of considering a two-way random-effects model, the individual and temporal effects satisfy the following, 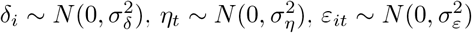 and

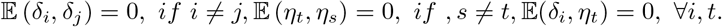 For the random effects models, there are three estimation methodologies for the parameters, which are available in the free R statistical software [25], through the plmtest command of the plm package. This command provides the following methods, “swar” [26] (default), “amemiya” [27], and “walhus” [28].
▪ One-way fixed-effects model The data come from 14 different regions, each with peculiar characteristics, which could suggest that the observations from the same region share some common characteristics, for example, climatic conditions, economic development, among other characteristics. Therefore, such characteristics could be related to the regressors, so it is advisable to incorporate the 14 regions as factors in the model and thus control by region. For these reasons, a model is proposed where the errors *u*_*it*_ consider only a time effect to control the contemporary correlation in the cross section as follows

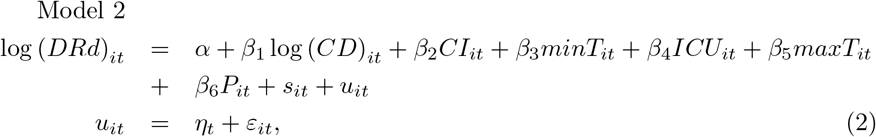

where *s*_*it*_ = *δ*_*i*_*R*_*i*_*I*_*i*_(*t*) with *R*_*i*_ represent the *i* th region and *I*_*i*_(*t*) 0, 1 an indicator variable, for *i* = 1, …, 14, *t* = 1, …, *T*, defined by

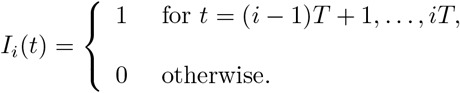

## 2. Results

In order to know the behavior of the variable of DRd in all the regions registered to priori and posterior of the pandemic, we consider the prior and posterior mean-variance tests, where the data from January 2020 are posterior to the pandemic and other months as prior to the pandemic. Table 2 reports the results obtained from applying the mean-variance tests. We have differentiated the cases of non-homogeneity of variance, in which case we use the Welch’s test. From the Table 2 we can say that deaths from respiratory diseases in all the study regions are significantly lower than before the outbreak, except in the Aysén region, which has a non-significant difference.

**Cuadro 2.** Mean-variance tests. This table reports the results of the mean-variance test of number of deaths due to respiratory diseases prior and posterior to the pandemic. The symbols ***, **, and * represent significance levels of 1 %, 5 %, and 10 %, respectively.

| Regions | Prior to the pandemic |  |  | Posterior to the pandemic |  |  | t-welch |
| --- | --- | --- | --- | --- | --- | --- | --- |
|  | obs | mean | variance | obs | mean | variance | mean(post.)-mean(prior) |
| Tarapacá | 27 | 13.29 | 24.60 | 11 | 10.36 | 12.25 | -2.93** |
| Antofagasta | 27 | 27.37 | 40.47 | 11 | 20.45 | 16.47 | -6.92*** |
| Coquimbo | 27 | 40.89 | 147.51 | 11 | 29.30 | 52.68 | -11.59*** |
| Valparaíso | 27 | 122.58 | 1374.09 | 11 | 80.17 | 326.15 | -42.41*** |
| Maule | 27 | 68.23 | 360.90 | 11 | 52.42 | 170.27 | -16.81*** |
| Biobío | 27 | 93.80 | 770.88 | 11 | 65.08 | 224.63 | -28.72*** |
| Araucanía | 27 | 71.19 | 266.08 | 11 | 46.75 | 87.47 | -24.44*** |
| Los Lagos | 27 | 49.67 | 207.69 | 11 | 35.36 | 22.65 | -12.30*** |
| Aysén | 27 | 4.12 | 5.73 | 11 | 3.82 | 2.57 | -0.29 |
| Magallanes | 27 | 8.89 | 11.49 | 11 | 5.19 | 2.76 | -3.71*** |
| Metropolitana | 27 | 437.38 | 21688.49 | 11 | 350.83 | 3688.88 | -86.55*** |
| Los Ríos | 27 | 22.96 | 61.19 | 11 | 15.82 | 28.56 | -7.14*** |
| Arica y P. | 27 | 10.18 | 11.62 | 11 | 7.18 | 7.16 | -3.00*** |
| Ñuble | 27 | 29.33 | 78.92 | 11 | 22.55 | 52.87 | -6.79** |

To complement the previous analysis, we consider one-way model with the post and pre-pandemic dummy variable to study its effect on Chile’s deaths from respiratory diseases, that is:

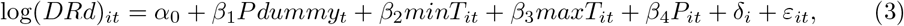

where *Pdummy*_*t*_ is a dummy variable, assuming a value of 1 for records after the COVID-19 pandemic and 0 for the months before the COVID-19 pandemic. This dummy variable can be used to directly compare the differences between the DRd before and after COVID-19. Among the models used to describe the relationship of equation (3) are the Pooling Model; which groups all the cross sections, without considering any form of individual effect and among the non-observable heterogeneity models are the Within Model, which is also called fixed effects models, and the random effects models defined above. Table (3) shows the estimated results of the aforementioned models. Note that in all models the *Pdummy*_*t*_ covariate is statistically significant. In particular, Column 4 shows that the COVID-19 pandemic has significantly reduced the DRd in a 34.12 %. It should be taken into consideration that, in the previous analysis, and as indicated in Eq (3), we only the regional fixed effect was controlled in the regression. Now let’s go a step further to consider panel data regression as Eqs (1)-(2). Next, the models proposed in this study (1 and 2) will be studied to analyze which are the variables that affect the DRd in Chile.

**Cuadro 3.** The individual effect model.

|  | OLS<br>pooling | Fixed effect<br>within | Random effect<br>swar | walhus | amemiya |
| --- | --- | --- | --- | --- | --- |
| *Pdummy* | -0.15310<br>(0.11084) | -0.34139***<br>(0.02846) | -0.34101***<br>(0.02890) | -0.34091***<br>(0.02910) | -0.34122***<br>(0.02848) |
| *maxT* | 0.10695***<br>(0.01212) | -0.01822*<br>(0.00883) | -0.01595<br>(0.00876) | -0.01542<br>(0.00877) | -0.01714*<br>(0.00874) |
| *minT* | -0.09122***<br>(0.01385) | -0.02976*<br>(0.01315) | -0.03257*<br>(0.01302) | -0.03322*<br>(0.01303) | -0.03111*<br>(0.01300) |
| *P* | 0.00115<br>(0.00099) | 0.00013<br>(0.00037) | 0.00019<br>(0.00037) | 0.00020<br>(0.00037) | 0.00016<br>(0.00037) |
| $R^2$ | 0.13782 | 0.35443 | 0.33965 | 0.33618 | 0.34741 |
| Adj. $R^2$ | 0.13128 | 0.33308 | 0.33464 | 0.33114 | 0.34245 |
| Num. obs. | 532 | 532 | 532 | 532 | 532 |
| $\sigma_\varepsilon$ | | | 0.29464 | 0.45444 | 0.29350 |
| $\sigma_\delta$ | | | 0.84821 | 1.17115 | 1.23975 |

\*\*\* $p < 0.001$ ; \*\* $p < 0.01$ ; \* $p < 0.05$

### 2.1. Using two-way fixed-effects model: Model 1

In this case, the performance of Model 1 was evaluated to determine the degree of impact of the pandemic on deaths from respiratory diseases in the regions of Chile. For this, it was controlled for both the regional fixed effect and the temporal effect in the panel data. Table (4) reports the benchmark estimation results of Model 1. From column 5, it shows that the number of deaths from Covid-19 has a significant negative effect on the response variable. On the other hand, the number of beds in ICU used by Covid-19 patients has a significant positive effect. Finally, among the climatic variables, the minimum temperature has a significant negative effect on the number of deaths from respiratory diseases in Chile, while precipitation has a positive effect. For example, for each percentage unit of increase in deaths from Covid-19, the number of deaths from respiratory diseases decreases by 7.6 %, while for each unit of increase in the number of ICU beds due to Covid-19 the deaths from respiratory diseases increased by 0.055 %. Another important aspect is that if the minimum temperature decreases by one degree the number of deaths from respiratory diseases rises 4.942 % and, a unit increase in the precipitation would result in a go up of 0.089 % in the number deaths from respiratory diseases. It is important to note that the performance of Model 1 with respect to adjusted *R*^2^ is low, which suggests that this model would not be the most appropriate to adjust the data under study.

**Cuadro 4.** The twoways effect model.

|  | OLS<br>pooling | Fixed effect<br>within | Random effect<br>walhus | amemiya | swar |
| --- | --- | --- | --- | --- | --- |
| $\log(CD)$ | 0.10598*<br>(0.04358) | -0.03090<br>(0.02171) | -0.05814**<br>(0.01817) | -0.05985***<br>(0.01760) | -0.07622***<br>(0.01807) |
| $CI$ | -0.01308***<br>(0.00220) | 0.00034<br>(0.00062) | 0.00016<br>(0.00063) | 0.00021<br>(0.00061) | -0.00037<br>(0.00077) |
| $ICU$ | 0.00255***<br>(0.00058) | 0.00030<br>(0.00017) | 0.00040*<br>(0.00017) | 0.00039*<br>(0.00016) | 0.00055**<br>(0.00020) |
| $maxT$ | 0.09466***<br>(0.01153) | 0.00543<br>(0.00877) | 0.00438<br>(0.00847) | 0.00185<br>(0.00836) | 0.01278<br>(0.00906) |
| $minT$ | -0.08177***<br>(0.01316) | -0.01078<br>(0.01489) | -0.02898*<br>(0.01329) | -0.02777*<br>(0.01330) | -0.04942***<br>(0.01304) |
| $P$ | 0.00071<br>(0.00093) | 0.00071<br>(0.00039) | 0.00078*<br>(0.00038) | 0.00073*<br>(0.00037) | 0.00089*<br>(0.00043) |
| $R^2$ | 0.23628 | 0.01495 | 0.04244 | 0.04704 | 0.09284 |
| Adj. $R^2$ | 0.22755 | -0.10118 | 0.03150 | 0.03615 | 0.08248 |
| Num. obs. | 532 | 532 | 532 | 532 | 532 |
| $\sigma_\varepsilon$ | | | 0.41075 | 0.25793 | 0.25955 |
| $\sigma_\delta$ | | | 1.05353 | 1.18026 | 0.24181 |
| $\sigma_\eta$ | | | 0.36376 | 0.22496 | 0.10289 |

\*\*\* $p < 0.001$ ; \*\* $p < 0.01$ ; \* $p < 0.05$

Figure 2 display the fitted values for each regression model of the data panel, where the red line represents the actual data, while the colored lines represent the fitted values for Model 1. From these figures, we can conclude that the best model that captures the trend structure is a Swar Type Random Effect Model.

**Figura 2.**
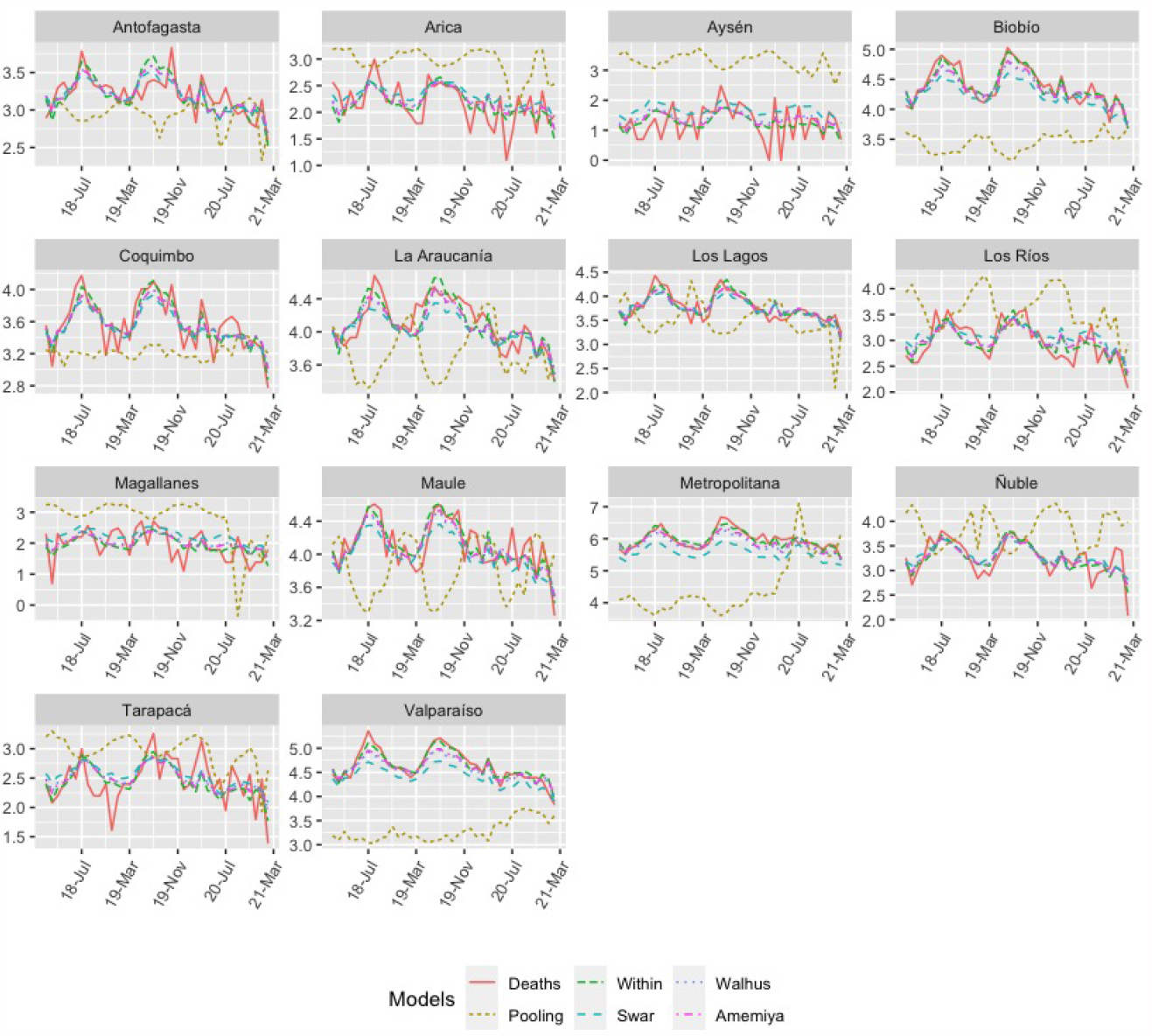
Model 1: Number of deaths from respiratory diseases (red lines) versus fitted values (colored lines).

### 2.2. Using one-way fixed-effects model: Model 2

Chile is one of the most centralized countries in Latin America and of the OCDE [30], that is, there is a high concentration of resources and development in the Metropolitan Region. Specifically, the capital of the country concentrates the best schools, hospitals, clinics, universities, cultural events and social development, among others. To eliminate the “centralization” effect, the regional effect was used as a covariate to study the impact of COVID-19 on the DRd based on the one-way fixed-effects model. Table (5) shows the estimated results with respect to Model 2. From Column 5, it is observed that COVID-19 has significantly reduced the number of deaths from respiratory diseases. For example, a unit percentage increase in deaths-Covid-19 would result in a 5.987 % drop in the number of deaths from respiratory diseases. On the other hand, a unit increase in the number of COVID-19 patients in intensive care leads to a 0.039 % increase in the number of deaths from respiratory diseases. Another important aspect is that if the minimum temperature increases by one degree, the number of deaths from respiratory diseases increases by 2,681 %. Unlike Model 1, the performance of Model 2 with respect to the adjusted *R*^2^ is high, this is supported by Fig 3 which shows the adjusted values of the panel regression model defined in Table (5). This suggests that Model 2 would be the most suitable for fitting the data in the study. A Hausman Type Test will determine whether individual effects should be treated as fixed or can be assumed incorrect with regressors, using a more efficient random effects specification. Under the hypothesis of no correlation between the regressors and the individual effects, this hypothesis, with a p-value of 18 %, is not rejected at the 5 % confidence level. Therefore, a random effect model with the Amemiya Method is more appropriate for this model, with an adjusted *R*^2^ = 0.95521. Finally, we can observe that the Regions of the extreme south (Region 11 and 12) and that of the extreme north (Region 15) show a decrease in the number of deaths due to respiratory diseases compared to the Tarapacá region (Region 1). While the rest of the regions present a significant increase in deaths from respiratory diseases when compared to the same region (Region 1). The foregoing is due to the fact that Regions 11, 12 and 15 have a lower number of population than the Tarapacá Region.

**Cuadro 5.**
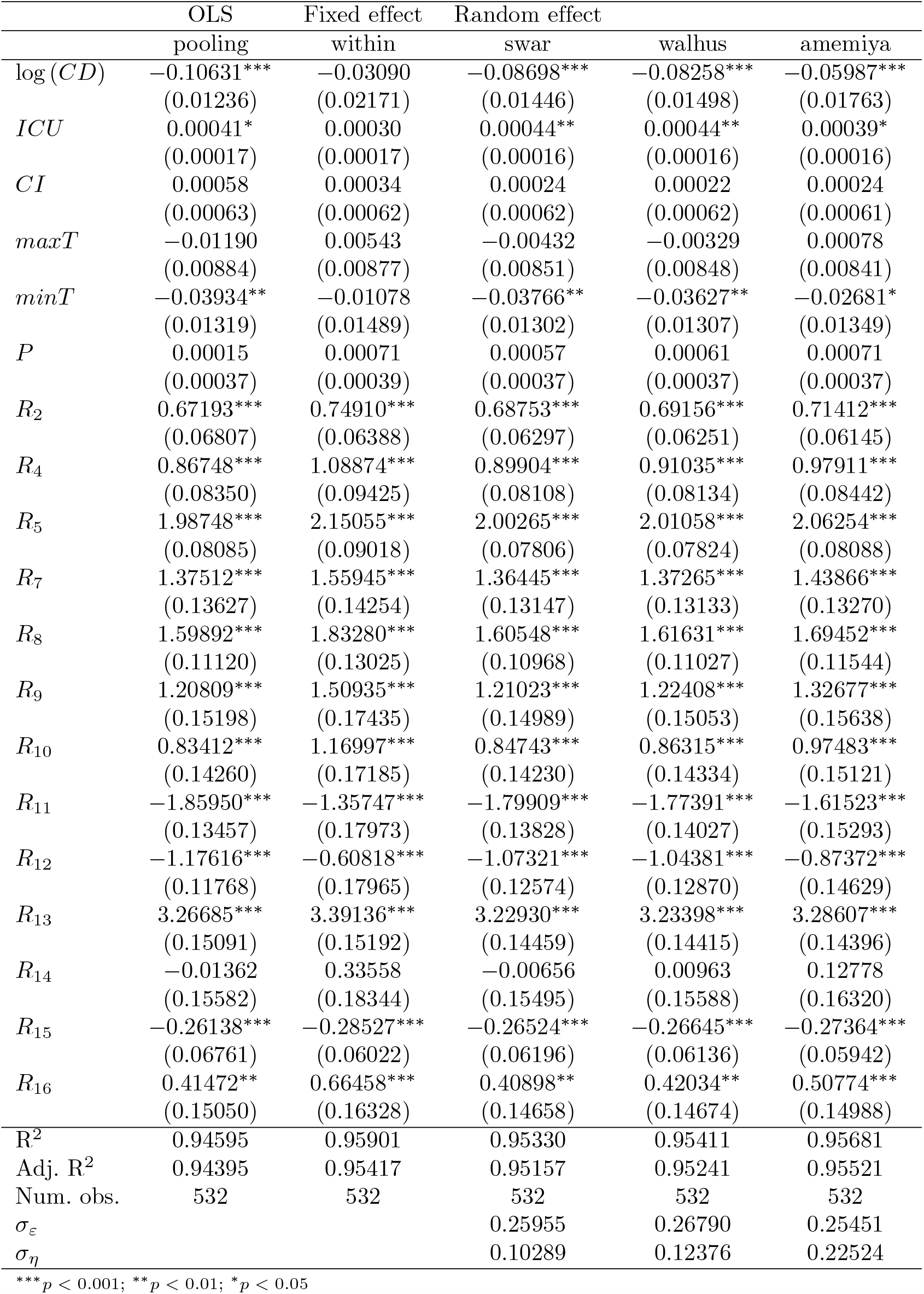
Time effect model.

|  | OLS | Fixed effect | Random effect |  |  |
| --- | --- | --- | --- | --- | --- |
|  | pooling | within | swar | walhus | amemiya |
| $\log(CD)$ | -0.10631***<br>(0.01236) | -0.03090<br>(0.02171) | -0.08698***<br>(0.01446) | -0.08258***<br>(0.01498) | -0.05987***<br>(0.01763) |
| $ICU$ | 0.00041*<br>(0.00017) | 0.00030<br>(0.00017) | 0.00044**<br>(0.00016) | 0.00044**<br>(0.00016) | 0.00039*<br>(0.00016) |
| $CI$ | 0.00058<br>(0.00063) | 0.00034<br>(0.00062) | 0.00024<br>(0.00062) | 0.00022<br>(0.00062) | 0.00024<br>(0.00061) |
| $maxT$ | -0.01190<br>(0.00884) | 0.00543<br>(0.00877) | -0.00432<br>(0.00851) | -0.00329<br>(0.00848) | 0.00078<br>(0.00841) |
| $minT$ | -0.03934**<br>(0.01319) | -0.01078<br>(0.01489) | -0.03766**<br>(0.01302) | -0.03627**<br>(0.01307) | -0.02681*<br>(0.01349) |
| $P$ | 0.00015<br>(0.00037) | 0.00071<br>(0.00039) | 0.00057<br>(0.00037) | 0.00061<br>(0.00037) | 0.00071<br>(0.00037) |
| $R_2$ | 0.67193***<br>(0.06807) | 0.74910***<br>(0.06388) | 0.68753***<br>(0.06297) | 0.69156***<br>(0.06251) | 0.71412***<br>(0.06145) |
| $R_4$ | 0.86748***<br>(0.08350) | 1.08874***<br>(0.09425) | 0.89904***<br>(0.08108) | 0.91035***<br>(0.08134) | 0.97911***<br>(0.08442) |
| $R_5$ | 1.98748***<br>(0.08085) | 2.15055***<br>(0.09018) | 2.00265***<br>(0.07806) | 2.01058***<br>(0.07824) | 2.06254***<br>(0.08088) |
| $R_7$ | 1.37512***<br>(0.13627) | 1.55945***<br>(0.14254) | 1.36445***<br>(0.13147) | 1.37265***<br>(0.13133) | 1.43866***<br>(0.13270) |
| $R_8$ | 1.59892***<br>(0.11120) | 1.83280***<br>(0.13025) | 1.60548***<br>(0.10968) | 1.61631***<br>(0.11027) | 1.69452***<br>(0.11544) |
| $R_9$ | 1.20809***<br>(0.15198) | 1.50935***<br>(0.17435) | 1.21023***<br>(0.14989) | 1.22408***<br>(0.15053) | 1.32677***<br>(0.15638) |
| $R_{10}$ | 0.83412***<br>(0.14260) | 1.16997***<br>(0.17185) | 0.84743***<br>(0.14230) | 0.86315***<br>(0.14334) | 0.97483***<br>(0.15121) |
| $R_{11}$ | -1.85950***<br>(0.13457) | -1.35747***<br>(0.17973) | -1.79909***<br>(0.13828) | -1.77391***<br>(0.14027) | -1.61523***<br>(0.15293) |
| $R_{12}$ | -1.17616***<br>(0.11768) | -0.60818***<br>(0.17965) | -1.07321***<br>(0.12574) | -1.04381***<br>(0.12870) | -0.87372***<br>(0.14629) |
| $R_{13}$ | 3.26685***<br>(0.15091) | 3.39136***<br>(0.15192) | 3.22930***<br>(0.14459) | 3.23398***<br>(0.14415) | 3.28607***<br>(0.14396) |
| $R_{14}$ | -0.01362<br>(0.15582) | 0.33558<br>(0.18344) | -0.00656<br>(0.15495) | 0.00963<br>(0.15588) | 0.12778<br>(0.16320) |
| $R_{15}$ | -0.26138***<br>(0.06761) | -0.28527***<br>(0.06022) | -0.26524***<br>(0.06196) | -0.26645***<br>(0.06136) | -0.27364***<br>(0.05942) |
| $R_{16}$ | 0.41472**<br>(0.15050) | 0.66458***<br>(0.16328) | 0.40898**<br>(0.14658) | 0.42034**<br>(0.14674) | 0.50774***<br>(0.14988) |
| $R^2$ | 0.94595 | 0.95901 | 0.95330 | 0.95411 | 0.95681 |
| Adj. $R^2$ | 0.94395 | 0.95417 | 0.95157 | 0.95241 | 0.95521 |
| Num. obs. | 532 | 532 | 532 | 532 | 532 |
| $\sigma_\varepsilon$ | | | 0.25955 | 0.26790 | 0.25451 |
| $\sigma_\eta$ | | | 0.10289 | 0.12376 | 0.22524 |

\*\*\* $p < 0.001$ ; \*\* $p < 0.01$ ; \* $p < 0.05$

**Figura 3.**
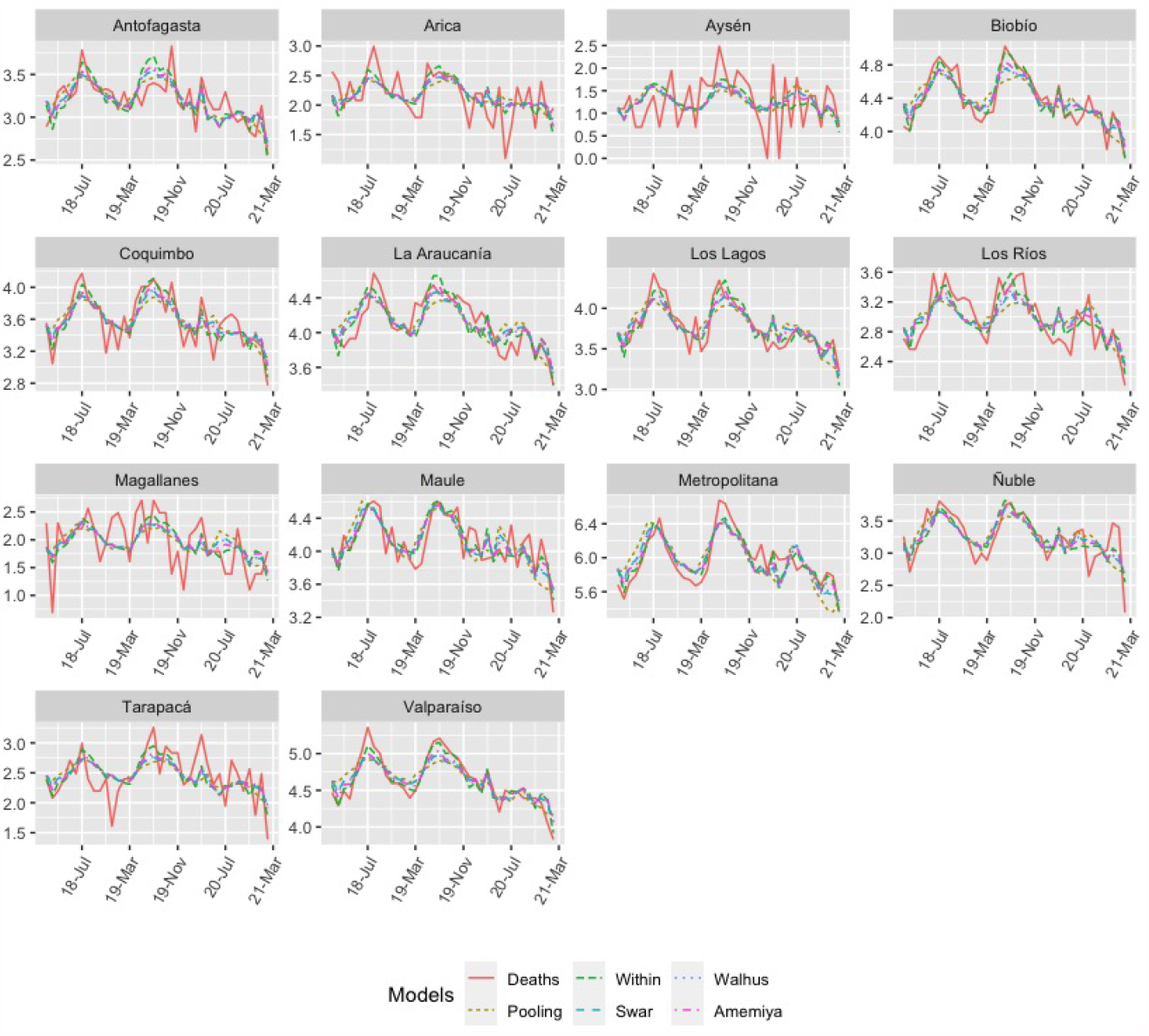
Model 2: Number of deaths from respiratory diseases (red lines) versus fitted values (colored lines).

## 3. Discussion

An important strength of this study is the general design used to assess the possible relationship between the COVID-19 pandemic with respect to climatic conditions and mortality from respiratory diseases. In all statistical analyzes performed in this study, the magnitude and direction of the observed phenomena were found to be consistent. Importantly, it was also revealed that by incorporating geographic location in Model 2 as an explanatory variable, the magnitude and significance of the observed results increased considerably.

Regarding the behavior of the variable number of deaths from respiratory causes (DRd) in all the regions registered after the pandemic, it was significantly lower than the records prior to the COVID-19 pandemic. These findings confirm the hypothesis of a study conducted in Brazil where the authors estimate an average underreporting of 40.68 % (range 25.9 % -62.7 %) for deaths related to COVID-19 in this country [8].

In this study it was also observed (Fig 3) that in the extreme North and South regions of Chile, Model 2 has a low performance to explain the number of deaths from respiratory diseases. Among the possible causes of the above, it can be mentioned that; the population density of these regions is lower compared to other areas. Another possible cause is the large difference in PIB per capita between the extreme regions of the country and the Metropolitan region with the rest of the regions of the national territory [30], which is associated with compliance with the different confinement measures, and the preventive health care that the inhabitants of these regions have had.

The findings of the present study have limitations. First, this study had a longitudinal type design. Therefore, in these types of studies, variables recorded both in time and by locality are needed, which are difficult to obtain. In this context, there are sociodemographic variables whose monthly records were not available by region, and therefore, they were not present in this study to analyze the association between COVID-19 and cultural differences to avoid deaths from respiratory diseases. It is proposed to carry out a subsequent study, which not only considers the geographical differences that affect self-protection habits against respiratory diseases and COVID-19, but also cultural differences.

Second, our findings could be affected by the number of individuals vaccinated against influenza during the COVID-19 pandemic. Another possible limitation of this study could be the existence of errors in the identification/recoding of the true cause of death in the database and errors in the recording/calculation of the climatic conditions analyzed in the database. It is presumed that, if such inaccuracies occurred in the data, they would have occurred with a similar frequency among the different databases examined. Therefore, if such phenomena were present in the data examined, the statistical power of this study would have been reduced. Still, the present study’s results provide some insight into the association between the COVID-19 pandemic and the number of deaths from respiratory diseases.

## 4. Conclusion

Our findings show that among the variables that affect the mortality rate from respiratory diseases are; the number of deaths from COVID-19 which has a negative effect, the number of patients with COVID-19 in the ICU, which has a positive effect, and the minimum temperature with a negative effect. These results are supported by the application of the panel regression for one-way random-effect model. We can conclude that the number of deaths from respiratory diseases in Chile decreased with the appearance of the COVID-19 pandemic.

## Data Availability

These variables 127 include the data on the number of deaths from respiratory diseases obtained from the 128 Ministry of Health https://deis.minsal.cl/, whereas the data on the variables 129 regarding the severity of COVId-19 were obtained from the Ministry of Science and 130 Technology https://www.minciencia.gob.cl/covid19. Finally, the data of the 131 temperature and precipitation variables were obtained from the updated report of the 132 Chilean Meteorological Directorate DGAC, which are available on the website 133 https://climatologia.meteochile.gob.cl.

## Referencias

1. Organización Panamericana de Salud. Actualización Epidemiológica Enfermedad por coronavirus (COVID-19). https://reliefweb.int/sites/reliefweb.int/files/resources/2021-mar-11-phe-actualizaci%C3%B3n-epi-COVID-19_0.pdf Accessed: 11 de marzo de 2021.

2. Ministerio de Salud de Chile. Departamento de Epidemiología. Informe epidemiológico. COVID-19. https://www.minsal.cl/wp-content/uploads/2021/03/Informe-epidemiolo%CC%81gico-102.pdf Accessed: 12-03-2021.

3. Ministerio de Salud de Chile. Departamento de Estadísticas e Información de Salud, DEIS. Informe semanal de defunciones por COVID-19 N°39 https://www.minsal.cl/wp-content/uploads/2021/03/Informe-Semanal-Defunciones-39.pdf Accessed: 11 de marzo de 2021.

4. Ministerio de Salud de Chile. Departamento de Estadísticas e Información de Salud, DEIS. Estadísticas de Defunciones por Causa Básica de Muerte. https://public.tableau.com/profile/deis4231#!/vizhome/DefuncionesSemanales1_0/DEF?publish=yes

5. Lallukka, Tea and Millear, Anoushka and Pain, Amanda and Cortinovis, Monica and Giussani, Giorgia. GBD 2015 mortality and causes of death collaborators. Global, regional, and national life expectancy, all-cause mortality, and cause-specifi c mortality for 249 causes of death, 1980-2015: a systematic analysis for the global burden of disease study 2015 (vol 388, pg 1459, 2016). Lancet. 2017; 389(10064):E1–E1.

6. Foro de las Sociedades Respiratorias Internacionales. (2017). El impacto mundial de la Enfermedad Respiratoria https://www.who.int/gard/publications/The_Global_Impact_of_Respiratory_Disease_ES.pdf

7. Ministerio de Salud de Chile. Departamento de Estadísticas e Información de Salud, DEIS. Estadísticas por Egresos Hospitalarios. Chile 2006, 2007, 2008 y 2009. http://www.deis.cl/estadisticasegresoshospitalarios/

8. Alonso, WJ and Schuck-Paim, C and Freitas, ARR and Kupek, E and Wuerzius, CR and Negro-Calduch, E and et.al. Covid-19 in context: comparison with monthly mortality from respiratory causes in each Brazilian state. Int Am J Med Health. 2020. https://iajmh.com/iajmh/article/view/93/107.

9. Shahzad, Khurram and Shahzad, Umer and Iqbal, Najaf and Shahzad, Farrukh and Fareed, Zeeshan. Effects of climatological parameters on the outbreak spread of COVID-19 in highly affected regions of Spain. Environmental Science and Pollution Research. 2020;27(31):39657–39666.

10. Jain, Mansi and Sharma Gagan Deep and Goyal, Meenu and Kaushal, Robin and Sethi, Monica. Econometric analysis of COVID-19 cases, deaths, and meteorological factors in South Asia. Environmental Science and Pollution Research. 2021: 1–17.

11. Briz-Redón, Álvaro and Serrano-Aroca, Ángel. The effect of climate on the spread of the COVID-19 pandemic: A review of findings, and statistical and modelling techniques. Progress in Physical Geography: Earth and Environment. 2020; 44(5):591–604.

12. Becchetti, Leonardo and Conzo, Gianluigi and Conzo, Pierluigi and Salustri, Francesco and others. Understanding the heterogeneity of COVID-19 deaths and contagions: the role of air pollution and lockdown decisions. Universitá degli studi di Torino, Department of Economics and Statistics. 2020.

13. Hashemi, Seyed A and Safamanesh, Saghar and Ghasemzadeh-moghaddam, Hamed and Ghafouri, Majid and Azimian, Amir. High prevalence of SARS-CoV-2 and influenza A virus (H1N1) coinfection in dead patients in Northeastern Iran. Journal of medical virology. 2021; 93(2):1008–1012.

14. Kim, David and Quinn, James and Pinsky, Benjamin and, Shah Nigam H and Brown, Ian. Rates of co-infection between SARS-CoV-2 and other respiratory pathogens. Jama. 2020; 323(20):2085–2086.

15. Luciano Kleber de Souza Luna and Dr. Ana Helena Perosa and Danielle Dias Conte and Joseane Mayara Almeida Carvalho and Vitória Rodrigues Guimarães Alves and Jessica Santiago Cruz and Nancy Bellei. Does COVID-19 infection impact on the trend of seasonal influenza infection? 11 countries and regions, from 2014 to 2020. International Journal of Infectious Diseases. 2020;97:1201–9712. https://doi.org/10.1016/j.ijid.2020.05.088.

16. Olsen, Sonja J and Azziz-Baumgartner, Eduardo and, Budd Alicia P and Brammer, Lynnette and Sullivan, Sheena and, Pineda Rodrigo F and Cohen, Cheryl and, Fry Alicia M. Decreased influenza activity during the COVID-19 pandemic—United States, Australia, Chile, and South Africa, 2020. American Journal of Transplantation. 2020;20(12):3681–3685.

17. Amato, Mauro and Werba JoséPablo and Frigerio, Beatrice and Coggi, Daniela and Sansaro, Daniela and Ravani, Alessio and Ferrante, Palma and Veglia, Fabrizio and Tremoli, Elena and Baldassarre, Damiano. Relationship between influenza vaccination coverage rate and COVID-19 outbreak: An Italian ecological study. Vaccines. 2020; 8(3):535.

18. Consortium, EBMPHET. COVID-19 Severity in Europe and the USA: Could the Seasonal Influenza Vaccination Play a Role?. Available at SSRN: http://dx.doi.org/10.2139/ssrn.3621446.

19. Petersen, Eskild and Koopmans, Marion and Go, Unyeong and, Hamer Davidson H and, Petrosillo Nicola and Castelli, Francesco and Storgaard, xMerete and Al Khalili, Sulien and Simonsen, Lone. Comparing SARS-CoV-2 with SARS-CoV and influenza pandemics.The Lancet infectious diseases. 2020. https://doi.org/10.1016/S1473-3099(20)30484-9.

20. Solomon DA, Sherman AC, Kanjilal S. Influenza in the COVID-19 Era. JAMA. 2020;324(13):1342–1343. doi:10.1001/jama.2020.14661.

21. Takahiro Itaya and Yuki Furuse and Kazuaki Jindai. Different patterns of Influenza A and B detected during early stages of COVID-19 in a university hospital in São Paulo, Brazil. Journal of Infection. 2020;81(2):e104–e105. https://doi.org/10.1016/j.jinf.2020.05.036.

22. World Health Organization Chronic respiratory diseases https://www.who.int/health-topics/chronic-respiratory-diseases#tab=tab_1

23. Gujarati, Damodar N. Basic Econometrics-Damodar N. Gujarati. McGraw-Hill. 2004.

24. Wooldridge, Jeffrey M. Econometric analysis of cross section and panel data. MIT press. 2010.

25. Team, R Core and others. R: A language and environment for statistical computing. Vienna, Austria. 2013.

26. Swamy, PAVB and, Arora Swarnjit S. The exact finite sample properties of the estimators of coefficients in the error components regression models. Econometrica: journal of the Econometric Society. 1972 : 261–275.

27. Amemiya, Takeshi. The estimation of the variances in a variance-components model. International Economic Review. 1971 : 1–13.

28. Wallace, T Dudley and Hussain, Ashiq. The use of error components models in combining cross section with time series data. Econometrica: Journal of the Econometric Society. 1969 : 55–72.

29. World Health Organization Coronavirus disease (COVID-19) advice for the public: when and how to use masks. https://www.who.int/emergencies/diseases/novel-coronavirus-2019/advice-for-public/when-and-how-to-use-masks

30. Mieres Brevis, Michelle. La dinamica de la desigualdad en Chile: Una mirada regional, Revista de análisis económico. 2020; 35(2):91–133.

